# Cost of hypertension illness and associated factors among patients attending hospitals in Southwest Shewa Zone, Oromia Regional State, Ethiopia

**DOI:** 10.1101/2019.12.17.19015198

**Authors:** Addisu Bogale, Teferi Daba, Dawit Wolde Daka

## Abstract

**Background:** Hypertension is a common vascular disease and the main risk factor for cardiovascular diseases. The impact of hypertension is on the rise in Ethiopia, so that, it is predictable that the cost of healthcare services will further increase in the future. We aimed to estimate the total cost of hypertension illness among patients attending hospitals in Southwest Shewa zone, Oromia Regional State, Ethiopia.

**Patients and methods:** Institution based cross-sectional study was conducted from July 1-30, 2018. All hypertensive patients who were on treatment and whose age was greater than eighteen years old were eligible for this study. The total cost of hypertension illness was estimated by summing up the direct and indirect costs. Bivariate and multivariate linear regression analysis was conducted to identify factors associated with hypertension costs of illness.

**Results:** Overall, the mean monthly total cost of hypertension illness was US $ 22.3 (95% CI, 21.3-23.3). Direct and indirect costs share 51% and 49% of the total cost, respectively. The mean total direct cost of hypertension illness per patient per month was US $11.39(95% CI, 10.6-12.1). Out of these, drugs accounted of a higher cost (31%) followed by food (25%). The mean total indirect cost per patient per month was US $10.89(95% CI, 10.4-11.4). Educational status, distance from hospital, the presence of companion and the stage of hypertension were predictors of the cost of illness of hypertension.

**Conclusion:** The cost of hypertension illness was very high when compared with the mean monthly income of the patients letting patients to catastrophic costs. Therefore, due attention should be given by the government to protect patients from financial hardships.

## Introduction

Hypertension is one of the main public health problem seen among adults worldwide. Hypertension is often defined as Systolic Blood Pressure (SBP) ≥140 mmHg or Diastolic Blood Pressure (DBP) ≥90 mmHg. It is the main risk factor to cardiovascular disease (CVD) such as heart diseases and stroke (1,2).

Worldwide, 45% of deaths as the result of heart diseases and 51% of deaths due to stroke was attributable to hypertension(3). In 2012 alone, 17.5 million people were died due to CVDs, which translated to every three people deaths in ten (4). Out of these deaths, over half, 9.4 million, deaths are caused by complications of hypertension(4,5). Hypertension was the fourth and seventh contributor to premature deaths in developed and developing countries, respectively (6).

The prevalence of hypertension is in a high rise wordwide over time (7) and the prevalence varies across the different regions and country income groups. Recent evidences indicated that an estimated 1.13 billion people have hypertension worldwide. Out of these, the majority (around two thirds) were located in low and middle income countries. The prevalence ranged from 27% in the World Health Organization (WHO) Africa region to 18% in the WHO region of Americans. The high rise in the prevalence of hypertension was attributed to the increase in the risk factors such as physical inactivity, the consumtion of tobacco and alcohol, unhealthy diet, overweight and obesity in those population groups (2,7).

In Ethiopia evidences showed that the prevalence and impact of non-communicable diseases (NCDs) including hypertension has increased over time(8). A meta analysis and other surveys conducted in different parts of Ethiopia revealed that the prevalence of hypertesion was increasing in Ethiopia. The prevalence was high and ranged from 24.4% to 34.7% (9–13). The latest WHO published data in 2017 indicated that deaths attributed to hypertension reached 11,050 deaths or 1.74% of the total deaths in Ethiopia. The age adjusted death rate in Ethiopia was 25.14 per 100,000 of population and this ranked Ethiopia 38^th^ in the world. Hypertension was ranked 11^th^ out of the top 20 causes of death in the country(14). Ethiopia was prone to double burden of diseases, both infectious and non-infectious diseses, that requires a urgent government actions(15).

The government of Ethiopia (GoE) has expressed its commitment for tacking premature deaths attributed to non-communicable diseases by signing the sustainable development goals (SDGs)(16) and incorporating related targets within the national health sector transformation plan (HSTP)(17). To effect this target, a comprehensive prevention and control strategic action plan of NCDs including hypertension was developed at national level with a particular emphasis to reducing risky behaviors(18).

Cost of health services is important determinant of the acceptability and utilization of health services by the society. The goal of health system is improving the health conditions of citizens through protecting financial catastrophes or hardships(19,20). The management of hypertension is cost intensive and hence, it may cause economic burden on citizens(21–24). Society may not afford available services leading to ineffectiveness of hypertension managements(25). Hypertension cost of illness was determined by various factors such as the presence of co-morbidity, stage of illness and distance to health facility(22,26). Therefore, it is crucial to quantify the cost of hypertension illness and identify the factors that determines the cost of hypertension illness among patients in Ethiopia.

Available investigations on the cost of hypertension illness in Ethiopia have focused on the direct costs of illness (26) and to our best knowledge little was known about the indirect costs of hypertension illness. Hence, the aim of this study was to estimate the cost of hypertension illness among patients in four hospitals of Southwest Shewa zone, Oromia Regional State, Ethiopia. We estimated the direct, indirect and total costs of hypertension illness. Moreover, we tried to identify the potential predictors of the total costs of hypertension illness.

## Patients and methods

### Study setting

The study was conducted in four hospitals of Southwest Shewa zone from August 13 to September 2, 2018. Southwest Shewa zone is one among the zonal administrations located in Oromia Regional State, 114 km far away from the capital city of Ethiopia, Addis Ababa. The total population of the zone was 1,101,129. Southwest Shewa zone comprised of 3 government hospitals, 1 other government organization (OGO) hospital, 54 health centers and 264 health posts. The hospitals were named as St. Luke general hospital; Tulo Bolo, Ameya and Bantu primary hospitals, respectively.

These hospitals provides high level curative services such as the management of non-communicable diseases like hypertension, diabetes, coronary heart diseases and stroke to the catchment population of the zone and neighboring zones.

### Study design and participants

Institution based cross-sectional study design was employed and the perspective of patients was used to estimate the cost of hypertension illness. All hypertensive patients who were on follow-up at hospitals and whose age was greater than 18 years old and who have been taken antihypertensive medications were eligible for this study. Pregnant women were excluded from this study to avoid gestational hypertension, as its effect may not last long.

Study sample size was determined using a single population mean formula(27). A previous report from Ethiopia public hospitals indicated that the mean or median drug prescription cost of hypertension was 100 Ethiopian Birr (ETB) or US $ 3.6 (SD 41.19 or 1.5)(26). By using the assumptions above, the estimated sample size at 95% confidence interval and 4% absolute allowable error (precision) yields 408 patients.

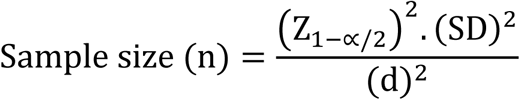

Where;

- Z1-α/2=is the value obtained from the standard normal distribution table for 100 (1-α) % confidence level and its value for 95% confidence level was 1.96.
- SD= is the population standard deviation (41.19)(26)
- d=is the absolute allowable error (or precision) in the estimation (4% of 100birr =4birr). The allowable error in estimating the average cost of prescription of drug for hypertension was 4 birr.

Though, as the number of hypertension patients on follow-up care (source population)) were less than 10,000 in the study area (N=1451), we corrected the sample size to the actual population size and this yields 318 patients. While, by adding 10% of non-response rate, the final estimated sample size yields 354 patients.

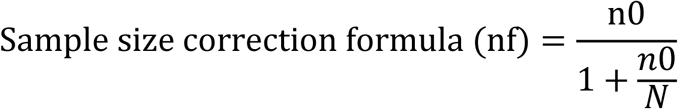

The calculated sample size was proportionally allocated to each of the study hospitals based on the caseload or hypertension patients per month. Accordingly, the number of patients per month and sample size allocated for each hospitals were: St. Luke General hospital (N=620, n=152); Tulu Bolo hospital (N=315, n=76); Amaya hospital (N=241, n=59); and Bantu hospital (N=275, n=67).

Each eligible patients who were presented to the study hospitals during the data collection period were approached and interviewed.

### Measurement

A structured data collection tools were used to gather data. The contents of the questionnaires were developed based WHO protocol for survey to determine costs of TB treatment program (28) and other related literatures(26).

The contents of the questionnaires were translated to local language (Afan Oromo) by an experienced translator and back translated to English with an independent translator for consistency. Five data collectors with a qualification of diploma nurse and who had also received additional training on the data collection questionnaires and research ethics were participated on data collection. The overall data collection processes were closely monitored and supervised by two health professionals who had a qualification of bachelor’s degree. Each eligible patients who visits the hospitals were approached and interviewed after explaining the aim of the study and obtaining a consent for participation.

### Techniques of cost estimation

The cost of hypertension illnesses analysis was conducted from the perspective of patients and a prevalence based model was employed to estimate cost(29). The main outcome variable was total cost of hypertension illness and it referred to the cost paid by patients as a consequence of a hypertension illness, typically it included both the direct and the indirect costs associated with hypertension illness. That means, total cost equals to the sum of direct and indirect cost of hypertension illness.

Direct cost referred to the opportunity cost of resources used for treating hypertension patients such as direct medical cost and direct non-medical cost. Direct medical cost was the amount of money that was paid for registration, laboratory investigation and medication or drugs in the study health facilities during the data collection period as presented below:

- Cost of hospital admission=Number of days admitted multiplied by cost per day
- Investigation cost=Number of investigations carried out multiplied by cost of each investigations (unit price)
- Cost of drug=Number of prescription given multiplied by cost of each drugs (unit price)

Direct non-medical costs referred to the amount of money that was paid for transportation, food, and accommodation for both patients and their caregiver (companion) to visit the hospital. Transportation costs were estimated based on the number of visits made to hospitals (round trips) to seek care and logging or accommodation costs were estimated based on the number of days of stay at hospitals as described below:

- Cost of transportation=Number of round trips from home to care provider multiplied by the cost of single trip
- Cost of lodging or accommodation=Money spent for food and accommodations multiplied by number of days patients and their companion were away from their home

Indirect cost was the value of resources lost due to a particular illness. Indirect costs were associated with loss of working time of the hypertension patients and their caregivers (companion). The loss of income of a person was due to absenteeism or missing a business appointment. Loss of income or earning was calculated for both employed and unemployed patients as shown below:

- 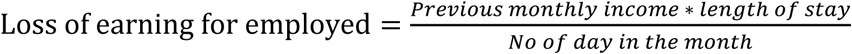
- 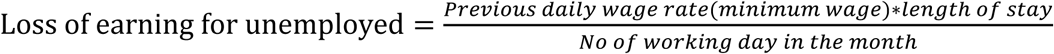

The costs of illnesses (both direct and indirect) were estimated as expressed for a 30-day period. The valuation of direct and indirect costs were based on a record review and patients recall.

The direct costs of hypertension illness were estimated by using micro-costing and bottom-up approaches(29). Accordingly, the healthcare consumptions were collected at individual patient level and the illness costs were also modeled at the same level. The direct costs such as costs of drugs, laboratory investigations and registrations were estimated based on the amount consumed by each patients. Health care consumption data were obtained through review of hospitals outpatient department (OPD) registration books and follow-up cards.

The indirect costs were estimated in terms of the productivity time losses of patients and their caregivers (companion). The human capital approach was used to value the productity time losses(29). Indirect costs were estimated for patients’ time lost from work during travel and consultation at care providers. First, the time forgone in seeking care and the productivity time lost were estimated. Then, the time forgone in seeking care and the productivity time lost were converted in to the daily indirect costs based on the daily wage rate. Finally, the value obtained by doing so was multiplied by the number of working days lost.

For monthly paid patients and companion, the daily wage rate was estimated by dividing their net monthly salary by 30 days. The income loses to patients who had no permanent job and farmers were estimated using local daily wage rate (Ethiopian birr per day) for unskilled labor and number of working days per month. The local daily wage rate for unskilled labor were obtained from the response of patients.

Whereas, the daily wage rate of daily paid patients and companion were estimated based on the patients reported daily earnings. The indirect costs of unemployed patients, such as students and patients who were not working due to physical or mental disability, were not considered in the calculations.

All direct costs, indirect costs and productivity losses were initially collected in Ethiopian currency (birr) and then converted to US $ during the analysis. A currency exchange rate of 1US $=27.471 Ethiopian Birr as of August 2018 was employed in the analysis.

## Data sources

The sources of information’s for estimating the costs of hypertension illness were both patients and medical records. Direct cost of illness was estimated based on the information obtained through patient interview and record review. Information related to indirect costs such as costs of transportation and logging were gathered through patients and companion interviews (Table 1).

**Table 1:** Cost item, data source and data type for cost of hypertension illness estimation among patients in South West Shewa Zone hospitals, Oromia Regional State, Ethiopia. August 2018

| Cost category | Data source | Method of data collection | Data type |
| --- | --- | --- | --- |
| Direct Medical Cost | Patient/health facility | Patient interview/record review | Cost of registration, drugs, laboratory, Admin cost ,if any investigations |
| Direct non-Medical Costs | Patient | Patient interview | Cost of transport for patients/Escort, lodging accommodation |
| Indirect Medical Cost | Patient | Patient interview | Income loss by patients and their families/relatives |

### Data analysis

Paper based data were checked for completeness and consistency. Following this, data were entered by using Epidata version 3.1 and exported to SPSS 20 for analysis.

The analysis was performed in such a way that, first, descriptive statistics was conducted to analyze information related to the characteristics of patients and to estimate the costs of hypertension illness. Following this, cost of illness data were checked for normality by using plots (Q-Q plots and histograms) and Kolmogorov-Smirnov test for normality (P>0.05). By doing so, the cost data were found to be right skewed and log transformation was performed to approximately conform the skewed data to normality.

Multivariate linear regression analysis was conducted to identify the predictors of the total cost of hypertension illness among patients.

In the regression model, independent variables with a probability value (P-value < 0.05) have been entered and only statistically significant variables (P-value < 0.05) were included into the final model and interpreted.

### Ethical Considerations

Research protocol was submitted to Jimma University Institute of health ethics review board and ethical clearance was obtained (protocol number IHRPGD/453/2018, July 2018). Research ethical clearance and permission was also obtained from Oromia regional health bureau and Southwest Shewa zone health department. The objective of the study was briefly explained to research participants followed by consent as assurance to their voluntary participation. The confidentiality of patient’s information was maintained throughout the data collection and analysis processes. Codes were used instead of patient identifying information’s and patient’s data were kept in a locked cabinet and in a password secured computers. Except the research team members, no one can access the patients data without due consent of individual patients.

## Results

### Participation

Out of the total planned 354 patients, 349 were participated in the study making a response rate of 98.6%.

### Characteristics of patients

Table 2 presented the characteristics of patients in the study area. The mean age of patients was 52 years (SD 14). The youngest patient was 22 years and the oldest one was 88 years. A bit more than half (52%) of patients were male. The majority of patients were married (80%), orthodox religion followers (56%), have no education (44%), employed (78%) and from urban settings (60%) (Table 2). The average monthly income of patients and households were 115.4 US $ (SD 70.1) and 136.8 US $ (SD 105.9), respectively.

**Table 2.** Characteristics of hypertensive patients at hospitals of Southwest Shewa zone, Oromia regional state, Ethiopia. August 2018

| **Variables** | **Observations**<br>**(n=349)** | **Mean/proportion (95% CI)** |
| --- | --- | --- |
| Age (mean± SD) | 51.2 (±13.9) | 51.2 (50-53) |
| Gender |  |  |
| **Male** | 181 | 52% (47-57%) |
| **Female** | 168 | 48% (43-53%) |
| **Total** |  |  |
| Marital status |  |  |
| **Married** | 281 | 80% (76-84%) |
| **Never married/single** | 27 | 8% (5-11%) |
| **Widowed/ Divorced** | 41 | 12% (9-16%) |
| Religion |  |  |
| **Orthodox** | 202 | 58% (53-63%) |
| **Muslim** | 78 | 22% (18-27%) |
| **Protestant** | 69 | 20% (16-24%) |
| Highest education level |  |  |
| **No education** | 154 | 44% (39-49%) |
| **Primary** | 57 | 16% (13-21%) |
| **Secondary** | 68 | 20% (16-24%) |
| **Higher** | 70 | 20% (16-25%) |
| Employment status |  |  |
| **Formal employment** | 144 | 41% (36-47%) |
| **Informal/unemployed** | 205 | 59% (54-64%) |
| Family size |  |  |
| **1-3** | 49 | 14% (11-18%) |
| **4-6** | 203 | 58% (53-63%) |
| **>6** | 97 | 28% (23-33%) |
| Residence |  |  |
| **Urban** | 210 | 60% (55-65%) |
| **Rural** | 139 | 40% (35-45%) |
| Average distance from hospital in (km) |  |  |
| **<10** | 169 | 48% (43-54%) |
| **11-20** | 57 | 16% (13-21%) |
| **21-30** | 42 | 12% (9-16%) |
| **31-40** | 36 | 11% (7-14%) |
| **41-50** | 20 | 6% (4-9%) |
| **50+** | 25 | 7% (5-10%) |
| Illness duration |  |  |
| **1-5 y** | 242 | 69% (64-74%) |
| **>5 y** | 107 | 31% (26-36%) |
| Stage of hypertension |  |  |
| **Prehypertension** | 113 | 32% (28-37%) |
| **Stage I** | 161 | 46% (41-51%) |
| **Stage II** | 75 | 22% (17-26%) |

### Cost of hypertension illness

#### Cost by patients

Table 3 presented the direct and indirect cost of hypertension illness among patients on follow-up care by cost category. The estimated total direct cost of hypertension illness was 3032.3 US $. The mean direct cost of hypertension illness was US $ 8.8 (95% CI, 8.2-9.3). Most (60%) of the direct costs of hypertension illness were attributed to medical costs (mean annual cost, US $ 5.2; 95% CI, 4.8-5.5). The direct medical costs comprised of cost for drugs, laboratory tests, registration and cost for non-prescribed drugs. Slightly over two-thirds (69%) of total medical costs were contributed by the cost of drugs alone (mean annual cost, US $ 3.6; 95% CI, 3.4-3.8). While costs for laboratory tests and registration were accounted of 13% (mean annual cost, US $ 1.2; 95% CI, 1.1-1.4) and 7% (mean annual cost, US $ 0.4; 95% CI, 0.37-0.39) of the total medical costs, respectively (Table 3).

**Table 3:** Patient direct and indirect costs of hypertension illness at hospitals of Southwest Shewa zone, Oromia region, Ethiopia. August 2018 (US $)

| **Cost item** | **Observations** | **Sum** | **Mean ± SD**<br>**(Median)** | **95% CI for**<br>**mean** |
| --- | --- | --- | --- | --- |
| **Total Direct Cost** | **349** | **3,032.252** | **8.8±4.9(7.6)** | **(8.2-9.3)** |
| **Direct medical cost** | **349** | **1,804.230** | **5.2±3.3(4.7)** | **(4.8-5.5)** |
| Cost of prescribed drugs | 349 | 1,247.752 | 3.6±1.9(3.3) | (3.4-3.8) |
| Cost of different laboratory tests | 186 | 231.371 | 1.2±0.9(0.9) | (1.1-1.4) |
| Cost registrations | 349 | 133.413 | 0.38±0.1(0.4) | (0.37-0.39) |
| Cost of non-prescribed drugs | 52 | 191.693 | 3.7±4.4(2.6) | (2.5-4.9) |
| **Direct non-medical cost** | **339** | **1,228.022** | **3.6±3.2(2.9)** | **(3.3-3.9)** |
| Cost of food during visit | 278 | 505.078 | 1.8±1.5(1.5) | (1.6-1.9) |
| Cost of transport | 302 | 369.335 | 1.2±1.2(0.7) | (1.1-1.4) |
| Cost of sport (gym) per month | 87 | 353.609 | 4.1±4.2(3.6) | (2.8-4.5) |
| **Income lost by patients** | **290** | **1,927.89** | **6.7±3.4** | **(6.3-7.0)** |
| Number of outpatient visit(in number) | 349 | 396 | 1.0±1.0 | (0.9-1.1) |
| Days absent from work | 349 | 459 | 1.3±1.3 | (1.2-1.5) |
| Travel time to the hospital (in minute) | 349 | 14,145 | 40.5±34.5 | (36.9-44.1) |
| Waiting time in the hospital (in minutes) | 349 | 10,715 | 30.7±14.7 | (29.2-32.2) |

Abbreviations: US \$, United States dollar; SD, standard deviation; CI, confidence interval

On average patients have visited hospitals once per month and half of patients have used bus as a means of transportation to visit hospitals. The mean lost earning day of patients due to hypertension illness was one working day. The total cost of lost earning was US $ 1, 927.9 and on average each patients paid US $ 6.7 (95% CI, 6.3-7.0) (Table 3).

#### Cost by companions

Table 4 presented the costs of hypertension illness paid by companions. Out of the total patients who have visited hospitals (349), the majority (87%; 95% CI, 83-90) had family or friend that accompany them. Half of the companions (52%; 95%, CI 46-57) have lost some portion of their income as they care patients. The mean direct cost of companion was US$ 3.1 (95% CI, 2.7-3.6) and the mean forgone earning by the companion as the result of hypertension illness was US $ 6.2 (95% CI, 5.9-6.5) (Table 4).

**Table 4:**
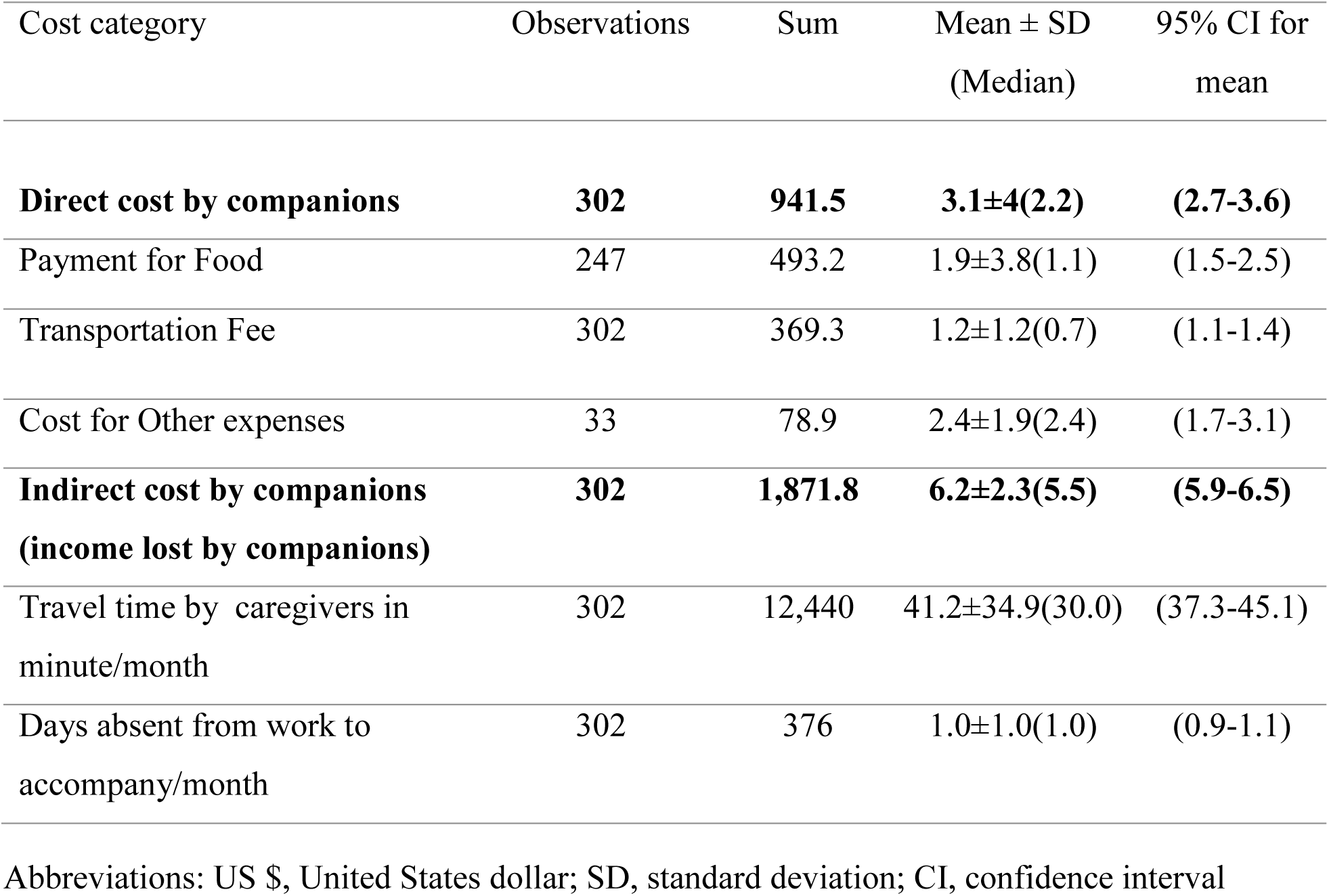
Companion direct and indirect costs of hypertension illness at hospitals of South West Shewa Zone, Oromia Regional State, Ethiopia. August 2018 (US $)

#### Total cost of hypertension illness

Table 5 presented the total cost of hypertension illness. The total cost of hypertension illness was US $ 7,773.5(mean monthly cost, US $ 22.3; 95% CI, 21.3-23.3). Out of these, direct costs accounted of 51% or US $ 3,973.8. The mean monthly and annual direct cost of hypertension illness per patient were US$ 11.4 (95% CI, 10.6-12.1) and US $ 136.6 (95% CI, 127.7-145.5), respectively (Table 5).

**Table 5:** Total cost of hypertension illness at hospitals of Southwest shewa zone, Oromia regional state, Ethiopia. August 2018 (US $)

| Cost category | Observations | Sum | Mean ± SD<br>(Median) | 95% CI for<br>mean |
| --- | --- | --- | --- | --- |
| **Total cost of hypertension** | **349** | **7,773.47** | **22.3±9.4(20.8)** | **(21.3-23.3)** |
| **Total direct cost of patients and companions** | **349** | **3,973.75** | **11.4±7.1(10.0)** | **(10.6-12.1)** |
| **Direct medical costs** | **349** | **1,804.23** | **5.2±3.4(4.7)** | **(4.8-5.5)** |
| Cost of prescribed drug | 349 | 1,247.752 | 3.6±1.9(3.3) | (3.4-3.8) |
| Cost of laboratory test | 186 | 231.371 | 1.2±0.9(0.9) | (1.1-1.4) |
| Cost of registration card | 349 | 133.413 | 0.4±0.1(0.4) | (0.37-0.39) |
| Cost non-prescribed drug | 52 | 191.693 | 3.7±4.4(2.6) | (2.5-4.9) |
| **Direct non-medical costs** | **339** | **2,169.52** | **6.5±5.9(5.2)** | **(5.9-7.1)** |
| Cost of Food | 299 | 998.25 | 3.3±4.2(2.2) | (2.9-3.8) |
| Cost of transport | 302 | 738.67 | 2.5±2.3(1.6) | (2.2-2.7) |
| Cost for sport(gym) | 87 | 353.61 | 4.2±4.2(3.6) | (3.3-5.0) |
| Other costs | 33 | 78.99 | 2.4±1.9(2.4) | (1.7-3.1) |
| **Indirect costs of patients and companions** |  |  |  |  |
| **Income lost by patients & companions** | **349** | **3,799.72** | **10.9±5.0(10.9)** | **(10.4-11.4)** |
| Travel time to the hospital in minute/month | 349 | 26,585.00 | 76.3±67.9(60.0) | (69.1-83.4) |
| Absent from works in days/month | 349 | 833.00 | 2.0±1.0(1.9) | (1.9-2.1) |

Abbreviations: US \$, United States dollar; SD, standard deviation; CI, confidence interval

The mean monthly and annual indirect costs of hypertension illness per patient were US $ 10.9 (95% CI, 10.4-11.4) and US $ 130.7, respectively. Indirect cost was the time devoted by hypertension patients and companion in one month recall period. It included detail time spent by patients and companion. Indirect cost was expressed by the lost days of patients and companion during the follow-up visits or seeking treatments. A total of 833 days were used by patients and companion with median of 1.9 days in one month period. The time spent by patients and companion to visit hospital was 26,585 minute with median of 60 minutes (round trip) (Table 5).

### Predictors of cost of hypertension illness

Table 6 presented the bivariate and multivariate linear regression analysis results. In the bivariate linear regression analysis variables such as gender, marital status, educational status, family size, residence, distance from hospital, presence of companion and stage of hypertension were associated with total cost of hypertension illness.

**Table 6:** Predictors of the total cost of hypertension illness among patients at hospitals of Southwest Shewa zone, Oromia regional state, Ethiopia. August 2018(US $)

| **Variables** | **Observations** | **Unadjusted $\beta$**<br>**coefficient (95% CI)** | **Adjusted $\beta$**<br>**coefficient (95% CI)** |
| --- | --- | --- | --- |
| Age in years (Mean ± SD) | 51.2 (±13.9) | 0.0001(-0.001,0.002) | |
| Gender |  |  |  |
| **Male** | 181 |  |  |
| **Female** | 168 | -0.070(-0.113, -0.027)* | -0.037(-0.077,-0.004) |
| Educational status |  |  |  |
| **No education** | 154 |  |  |
| **Primary** | 57 | -0.068(-0.126,-0.009)* | -0.072(-0.124,-0.020)* |
| **Secondary** | 68 | 0.007(-0.047,0.062) |  |
| **Tertiary** | 70 | 0.002(-0.052,0.056) |  |
| Marital status |  |  |  |
| **Married** | 281 |  |  |
| **Single** | 27 | 0.030(-0.051,0.112) |  |
| **Divorced/widowed** | 41 | -0.091(-0.157,-0.024)* | -0.057(-0.121,0.006) |
| Family size |  |  |  |
| **1-3** | 49 |  |  |
| **4-6** | 203 | 0.106(0.044,0.169)* | 0.029(-0.010,0.068) |
| **>6** | 97 | 0.122(0.050,0.195) |  |
| Residence |  |  |  |
| **Urban** | 210 |  |  |
| **Rural** | 139 | 0.062(0.018,0.106)* | 0.020(-0.021,0.060) |
| Distance from hospital (Km) | 21.6±23.7 | 0.003(0.002,0.004)* | 0.003(0.002,0.004)* |
| Presence of companion |  |  |  |
| **Yes** | 153 | 0.106(0.064-0.149)* | 0.096(0.057,0.135)* |
| **No** | 196 |  |  |
| Stage of hypertension |  |  |  |
| **Pre-hypertension** | 113 |  |  |
| **Stage one** | 161 | -0.007(-0.051,0.036) |  |
| **Stage two** | 75 | 0.074(0.021,0.126)* | 0.070(0.023,0.118)* |
| Duration of hypertension illness |  |  |  |
| **1-5 year** | 242 |  |  |
| **>5 year** | 107 | -0.027(-0.074,0.020) |  |
| Presence of comorbidity |  |  |  |
| **Yes** | 227 |  |  |
| **No** | 122 | 0.012(-0.034,0.057) |  |

Abbreviations: US \$, United States dollar; SD, standard deviation; CI, confidence interval;
\*P-value<0.05

After adjusted for other independent variables, educational status, distance from hospital, presence of companion and stage of hypertension were statistically associated with the total cost of illness. Twenty three percent (23%) of the variability in the total cost of hypertension illness was explained by these variables (Table 6).

Patients with no education have a higher total cost of hypertension illness compared to patients with a primary educational level (β-coefficient=-0.07; 95% CI, (−0.124,-0.020).

The total cost of hypertension illness increased with the increase in distance to hospital. With a unit increase in distance to hospitals, the total cost of hypertension illness increased by about US $ 0.003(95% CI, 0.002, 0.004) keeping the other variables constant.

Moreover, the total cost of hypertension illness was associated with the presence of companion and the stage of hypertension. The total cost of hypertension was higher among patients who have companion compared to those who didn’t have (β-coefficient=0.096; 95% CI, 0.057, 0.135). Likewise, the total cost of hypertension illness was higher among patients in the hypertension stage of two compared to patients in the prehypertension stage (β-coefficient=0.070; 95% CI, 0.023, 0.118) (Table 6).

## Discussion

Cost of illnesses analysis is useful to understand the burden of a particular disease in a society(29–31). In this study we estimated the cost of hypertension illness among patients on follow-up care. The estimated total cost of hypertension illness was 7775.5 USD and out of these direct cost accounted of 51%. The average annual cost of hypertension illness among patients was US $ 267.2. Likewise, the annual mean direct and indirect costs per patients were US $ 136.6 and US $ 130.7, respectively. In this study variables such as educational status, distance from hospital, the presence of companion and stage of hypertension were the predictors of the total cost of hypertension illness in the study area.

The cost of hypertension illness was lower that a study conducted in Kenya, where the mean annual direct and indirect cost to patient were US$ 304.8 (95% CI, 235.7‐374.0) and US$ 171.7 (95% CI, 152.8‐190.5), respectively (23). However, the cost of hypertension illness was higher than a study conducted in India (32) and Addis Ababa (26). This difference might be due to variation in settings, period and methods of cost estimation employed.

A bit over half of the total cost of hypertension illness was contributed by direct medical and direct non-medical costs. Most of the direct costs were due to drugs followed by accommodation and transportation. In the study setting, direct medical cost accounted of 29% of the direct cost of hypertension illness. A comparable result was also reported in other settings in which much of the cost of hypertension illness was attributed to medicines(21–23,26,32,33). This might be related to the high cost of hypertension drugs and this implied a higher rates of catastrophic costs on hypertension patients.

Nonetheless, the indirect costs of hypertension illness were also equally important. These costs included the time spent by patients and their companion, and expressed as lost work days. The majority of patients had companion and the mean forgone earning by the companion due to hypertension illness was US$ 6.2. The estimated average annual indirect cost of hypertensive illness was US $ 130.7 and this is lower than a report from Kenya(23). However, the indirect cost of hypertensive illness was high in the study area and it accounted of 49% of the total cost of illness. Indirect cost of hypertensive illness was attributed to the travel time to the hospital and absent from work. This indicates the need for expanding hypertension management centers and enhancing the control of hypertension in a community. In Ethiopia the coverage of non-communicable diseases management services was low and increasing steadily (34).

The cost of hypertension illness was determined by educational status, distance from the hospital, presence of companion and the stage of hypertension illness. The cost of hypertension illness was higher in no educated patients than the primary level education patients. This might be due to the reason that in the study setting most of the patients with a no educational status were at stage two of hypertension and the complications related to hypertension may let them to cost more. The total cost of hypertension was also associated with the presence of companion and distance to hospital. With a unit increase in distance to hospital, the cost of hypertension illness increased by about 0.003 US $. This is due to the fact that the companion consumption (food and transport) and the transportation cost of patients to visit hospitals may increases the total cost of hypertension illness. Likewise, patients who were in stage two of hypertension have a higher cost of illness than those in the prehypertension stage. This implies that the complications of hypertension is a significant contributor to increase in the cost of illness and this warrants the early diagnosis and management of hypertension. Our findings are comparable to other reports elsewhere(21,32).

Though our study have a many strengths, it has also some limitations. The cost of illness analysis was limited to patient’s perspective and doesn’t addressed other costs of the health system (such as costs related to the provider, family and society at large). Moreover, the intangible cost (costs of pain, suffering, stigma and discrimination) associated with hypertensive illness were not also addressed in our study.

## Conclusion

Overall, this study have revealed that the total cost of hypertension illness was high in the study area. The mean total cost of hypertension illness was high compared to the mean monthly income of patients and households, letting patients to catastrophic costs. Unless urgent measures were taken, the increased prevalence of hypertension combined to the high cost of hypertension management in Ethiopia may bare individuals to financial hardships. Thus, the government should take attention in preventing further catastrophic costs resulted from the complications and mortality of hypertension.

## Data Availability

Research data were available and can be accessed from the indicated addresses

## Acknowledgments

Our sincere gratitude should goes to the participants of the study and the administrative offices for their cooperation throughout the data collection process.

## Author contributions

All authors contributed toward conceiving idea, study design and data analysis. DWD drafted the manuscript and AB and TB revised the paper. All authors also gave final approval of the paper.

